# Predictors of upper respiratory *Streptococcus pneumoniae* colonization among working-age adults with prevalent exposure to overcrowding

**DOI:** 10.1101/2024.01.07.24300941

**Authors:** Anna M. Parker, Nicole Jackson, Shevya Awasthi, Hanna Kim, Tess Alwan, Anne L. Wyllie, Katherine Kogut, Nina Holland, Ana M. Mora, Brenda Eskenazi, Lee W. Riley, Joseph A. Lewnard

**Author notes:** Corresponding author: Joseph A. Lewnard, 2121 Berkeley Way, Room 5410, Berkeley, California 94720, United States.

## Abstract

**Background:** The epidemiology of adult pneumococcal carriage remains poorly understood. We assessed risk factors for pneumococcal carriage among adults in socioeconomically-disadvantaged farmworker communities with prevalent overcrowding.

**Methods:** From July-November 2020, we administered surveys and collected saliva from working-age adults within clinical and community settings throughout Monterey County, California. We detected pneumococci via qPCR assays targeting *lytA* and *piaB* genes. We evaluated predictors of pneumococcal carriage detection via conditional logistic regression.

**Results:** Among 1,283 participants, 117 (9.1%) had any detectable pneumococcal carriage and 53 (4.1%) carried pneumococci at a higher density threshold (*lytA* and *piaB* c*_T_<*35). Pneumococcal carriage was more common among individuals of lower socioeconomic status, as signified by indigenous background (odds ratio [OR]=3.94 [95% confidence interval: 2.36-6.60]), engagement in fieldwork (OR=2.01 [1.12-3.63]), and lacking high school-level education (OR=0.48 [0.26-0.90]). Within the household, carriers were more likely than non-carriers to be exposed to children aged <5 years (OR=1.45 [0.95-2.20]), and to be exposed to crowding (OR=1.48 [0.96-2.30] and 2.84 [1.20-6.73], respectively, for participants living in households with >2-4 and >4 persons per bedroom vs. ≤2 persons per bedroom). Household crowding was independently associated with increased risk of carriage among participants not exposed to children aged <5 years (OR=2.05 [1.18-3.59] for participants living in households with >2 vs. ≤2 persons per bedroom). Exposure to children aged <5 years and exposure to household crowding were each associated with increased pneumococcal density among carriers (*piaB* c*_T_* difference of 2.04 [0.36-3.73] and 2.44 [0.80-4.11], respectively).

**Conclusions:** While exposure to young children was an important risk factor for pneumococcal carriage, the association of crowding with increased risk of carriage in households without young children suggests transmission among adults may also occur in crowded congregate settings.

## INTRODUCTION

The bacterial pathogen *Streptococcus pneumoniae* (pneumococcus) is a prominent cause of severe invasive infections as well as mucosal conditions such as non-bacteremic pneumonia among both children and adults [1,2]. Commensal carriage of pneumococci in the upper respiratory tract is the source of transmission and a necessary precursor to pneumococcal diseases [3]. Children aged 2-4 years are widely understood to serve as the primary reservoir for pneumococcal transmission, with pediatric carriage prevalence estimates spanning 17-90% across various settings [4–6].

Whereas adults account for a majority of pneumococcal disease burden in the United States (US) [7,8], and may contribute to transmission across age groups, understanding of the prevalence and predictors of adult carriage remains limited [9]. Studies employing conventional nasopharyngeal sampling techniques and culture-based identification of pneumococci have yielded low (<5%) estimates of carriage prevalence among adults, although novel molecular approaches enabling pneumococcal carriage detection from saliva have identified ≥20% prevalence of adult carriage in some populations [10]. Studies employing these higher-sensitivity methods for sampling and detection offer an opportunity to better understand pneumococcal epidemiology among adults, providing insight into risk factors for carriage acquisition and the role of adults in transmission within distinct communities.

Working-age adults exposed to crowded conditions have emerged as an important risk group for pneumococcal disease, with invasive pneumococcal disease outbreaks reported among miners [11], military recruits [12], shipyard workers [13], prisoners [14], displaced migrants [15], residents of homeless shelters [16], and others living or working within close, congregate settings [17]. These outbreaks suggest that pneumococcal transmission may occur among adults under crowded conditions without primary involvement of young children. In the US, agricultural workers may comprise an at-risk group for such transmission due to prevalent exposure to overcrowding, poor ventilation, and suboptimal hygienic conditions in their housing, transportation, and working environments [18]. As part of a prior cross-sectional study among adult farmworkers within California’s Salinas Valley, we collected saliva samples for pneumococcal carriage detection while administering SARS-CoV-2 testing in both clinical and community settings from July to November, 2020 [19,20]. We revisited data from this study aiming to characterize risk factors and serotypes associated with adult pneumococcal carriage.

## METHODS

### Study setting

The Salinas Valley in Monterey County, California is home to an agricultural workforce of approximately 50,000 resident workers and 40,000 seasonal workers [18]. Most are undocumented Latino migrants from Mexico and Central American countries, who often reside in crowded multigenerational housing, including in makeshift residences [21,22]. The local workforce also includes holders of temporary agricultural H-2A visas, many of whom receive employer-provided group housing within motels or dormitories [23]. We undertook the parent study in partnership with Clínica de Salud del Valle de Salinas (CSVS), a Federally-Qualified Health Center focused on providing health care for agricultural workers and their families within the region. At the time of our study, CSVS offered SARS-CoV-2 testing at no cost to all members of the community, both at their network of Salinas Valley clinics and off-site at community testing events held at housing complexes, agricultural fields, community health fairs, and other sites.

### Study design

Enrollment for the study occurred between July 16^th^ and November 30^th^, 2020. For the primary study, we invited non-pregnant farmworkers aged ≥18 years who spoke English or Spanish and who received SARS-CoV-2 testing via CSVS at clinical and non-clinical settings to participate in a study on risk factors for SARS-CoV-2 infection within the farmworker community. To expand the study sample for analyses specifically focused on pneumococcal carriage and SARS-CoV-2 infection [20], we also invited adults of any occupational status, who were not pregnant women, to complete an abbreviated version of the study procedures. Regardless of the version of the study into which they were enrolled, all participants provided a ∼3mL saliva sample at the site where they completed their SARS-CoV-2 testing with CSVS (whether in clinical or non-clinical settings) and responded to a questionnaire addressing sociodemographic characteristics, household, and workplace exposures, and clinical symptoms. After collection, untreated saliva samples were frozen at –20°C immediately and thereafter stored at –80°C until delivery to the laboratory for processing. The study protocol was approved by the Office for the Protection of Human Subjects at University of California, Berkeley.

### Molecular detection

We used previously-validated protocols [24] for molecular detection of pneumococcal carriage in saliva samples after a culture-enrichment step in which saliva samples were incubated on trypticase soy agar supplemented with 7% sheep’s blood and 5 mg/L gentamicin (**Supplementary Methods**). After bacterial DNA isolation, we evaluated pneumococcal carriage based on the presence of *lytA* and *piaB* genes via quantitative polymerase chain reaction (qPCR). We defined adults with *lytA* and *piaB* cycle threshold (c*_T_*) values <40 as pneumococcal carriers; those with c*_T_* values <35 for both *lytA* and *piaB* were considered to carry pneumococci at higher density. Adults with *lytA* c*_T_* values <40 but *piaB* c*_T_* values ≥40 were considered probable carriers of other *lytA*-positive oral Streptococci [25].

We accounted for SARS-CoV-2 infection status within statistical analyses (as described below) to correct for a potential synergistic relationship between SARS-CoV-2 and pneumococci, as identified within the primary study [20]. To identify SARS-CoV-2 infection, oropharyngeal samples (collected at the same visit as study-related saliva samples) were tested via the qualitative Aptima nucleic acid transcription-mediated amplification assay (Hologic, Marlborough, Massachusetts, US).

### Serotyping

We conducted molecular serotyping using DNA from bacterial culture-enriched samples via a previously-validated quadruplex qPCR technique [26]. To overcome the low specificity of serotyping results from colony-free detection methods (e.g., due to presence of other bacteria encoding the serotype-specific *cps* genes), we included only results where serotype-specific c*_T_* values fell within a range of ±2 relative to the *piaB* c*_T_* value. We excluded detections of serotypes 4, 9V/9A, 17F, 21, 23A, 33A/33F and 35B due to prior evidence of low assay specificity for these serotypes [24,27].

### Statistical analyses

We compared the distribution of risk factors among individuals according to pneumococcal carriage status. Risk factors included age, sex, country of birth, language spoken, household income, marital status, cigarette smoking, years residing in the US, H-2A visa status, educational attainment, exposure to children at home, household size and crowding, access to in-home laundry facilities, engagement in field work, use of face coverings when outside the home, and regular handwashing after touching objects outside the home. We calculated unadjusted odds ratios (ORs) using conditional logistic regression, matching participants on recruitment venue (clinical or outreach testing site) and concurrent SARS-CoV-2 infection (positive or negative) via regression strata. We used the Akaike Information Criterion to identify variables for inclusion in conditional logistic regression models estimating adjusted odds ratios (aORs) for these exposures, again defining strata for recruitment venue and concurrent SARS-CoV-2 infection. Individuals without pneumococcal carriage provided the reference group for analyses of pneumococcal carriage or higher-density pneumococcal carriage outcomes.

We conducted multiple imputation with five pseudo-datasets to accommodate missing exposure data and pooled results across datasets via Rubin’s rules [28]. As a sensitivity analysis, we also fit models defining SARS-CoV-2 as a covariate rather than a stratum (matching) variable. We further conducted analyses limiting the study population to individuals with negative SARS-CoV-2 testing results (*n*=1,134 [88.4% of 1,283 otherwise-eligible participants]) and individuals without missing exposure data (*n*=1,270 [99.0% of 1,283]).

Because higher carriage density may indicate recent acquisition of pneumococcal carriage [29], we also explored associations of the risk factors listed above with carriage density in order to better understand potential sources of transmission. We used linear regression models to measure adjusted associations of each covariate with c*_T_* values for *piaB* among individuals found to carry pneumococcus; we used *piaB* as the primary target for measuring carriage density due to greater specificity as an indicator of pneumococcal carriage. The resulting effect measure can be interpreted as the adjusted mean difference in c*_T_* values when comparing pneumococcal carriers across exposure categories. We also report findings with respect to c*_T_* values for *lytA* as a supplemental analysis, noting that this measure may relate to density of other *lytA*-positive oral Streptococci, even among pneumococcal carriers, due to its lower specificity.

Last, we aimed to test whether pneumococcal carriage was associated with presence of respiratory symptoms. We limited these analyses to individuals with negative SARS-CoV-2 testing results; potential associations between symptoms and pneumococcal interactions with SARS-CoV-2 infection were addressed in prior analyses [20]. We defined individuals experiencing respiratory symptoms as those who reported nonproductive cough, productive cough, blocked nose, runny nose, sneezing, hoarseness, a tickling sensation in the throat, sore throat, painful sinuses, difficulty breathing, wheezing, or shortness of breath. We used conditional logistic regression models, stratified on recruitment clinical or outreach recruitment venue, to calculate ORs for outcomes of pneumococcal carriage and higher-density pneumococcal carriage. We also used linear regression to evaluate the mean difference in *lytA* and *piaB* c*_T_* values among pneumococcal carriers who experienced or did not experience symptoms, consistent with the density analyses described above.

We conducted analyses using R (version 4.1.1; R Foundation for Statistical Computing, Vienna, Austria).

## RESULTS

### Enrollment and inclusion

Of 1,309 participants who enrolled in the study, our analyses included 1,283 (1,099 farmworkers within the primary study group and 184 additional individuals who participated in the abridged supplemental study not restricted to farmworkers). We excluded 8 participants who did not meet eligibility criteria, 3 participants who did not provide a saliva sample, and 15 participants whose saliva specimens did not meet quality criteria (for reasons including label mismatch, risk of contamination, or insufficient saliva volume). Among participants included, 566 (44.1% of 1,283) enrolled in clinical settings and 717 (55.9%) enrolled in outreach settings (**Table 1**; **Table S1**). Most participants spoke Spanish at home (1,055/1,283; 82.2%), never attended high school (774/1,282; 60.4%), and lived in households earning <$25,000 annually (622/1,212; 51.3%). As compared to participants recruited in clinical settings, a greater proportion of those recruited via outreach testing were born in the US, had completed high school, lived in apartments or hotels (versus other housing types), and were not infected with SARS-CoV-2.

**Table 1.** Characteristics of the study population, by recruitment.

| Characteristics |  | Participants, no. (%) |  |  |
| --- | --- | --- | --- | --- |
|  |  | All | Clinic sample | Outreach sample |
|  |  | N=1,283 | N=566 | N=717 |
| Study population |  |  |  |  |
|  | Primary study | 1,099 (85.7) | 551 (97.3) | 548 (76.4) |
|  | Supplemental study | 184 (14.3) | 15 (2.7) | 169 (23.6) |
| Age range, years |  |  |  |  |
|  | 18-29 | 344 (26.8) | 145 (25.6) | 199 (27.7) |
|  | 30-39 | 306 (23.9) | 137 (24.2) | 169 (23.6) |
|  | 40-49 | 327 (25.5) | 158 (27.9) | 169 (23.6) |
|  | ≥50 | 306 (23.9) | 126 (22.2) | 180 (25.1) |
| Sex |  |  |  |  |
|  | Female | 698 (54.4) | 304 (53.7) | 394 (55.0) |
|  | Male | 585 (45.6) | 262 (46.3) | 323 (45.0) |
| Country of birth |  |  |  |  |
|  | Mexico | 1,012 (78.9) | 484 (85.5) | 528 (73.6) |
|  | United States | 226 (17.6) | 54 (9.5) | 172 (24.0) |
|  | Other | 45 (3.5) | 28 (4.9) | 17 (2.4) |
| Language spoken at home |  |  |  |  |
|  | Spanish | 1,055 (82.2) | 457 (80.7) | 598 (83.4) |
|  | English | 118 (9.2) | 25 (4.4) | 93 (13.0) |
|  | Indigenous language<sup>2</sup> | 110 (8.6) | 84 (14.8) | 26 (3.6) |
| Educational attainment (N=1,282) |  |  |  |  |
|  | Never attended school | 63 (4.9) | 48 (8.5) | 15 (2.1) |
|  | Some primary school | 460 (35.9) | 225 (39.8) | 235 (32.8) |
|  | Primary school completed | 251 (19.6) | 117 (20.7) | 134 (18.7) |
|  | Some high school | 148 (11.5) | 66 (11.7) | 82 (11.4) |
|  | High school completed | 360 (28.1) | 109 (19.3) | 251 (35.0) |
| Annual household income (N=1,212) |  |  |  |  |
|  | <25,000 | 622 (51.3) | 286 (53.5) | 336 (49.6) |
|  | 25,000-34,999 | 284 (23.4) | 113 (21.1) | 171 (25.3) |
|  | 35,000-49,000 | 195 (16.1) | 88 (16.4) | 107 (15.8) |
|  | ≥ 50,000 | 111 (9.2) | 48 (9.0) | 63 (9.3) |
| Washing machine in residence (N=1,099)<sup>1</sup> |  |  |  |  |
|  | No | 408 (37.1) | 214 (38.8) | 194 (35.4) |
|  | Yes | 691 (62.9) | 337 (61.2) | 354 (64.6) |
| H-2A visa (N=946)<sup>1</sup> |  |  |  |  |
|  | No | 881 (93.1) | 477 (96.0) | 404 (90.0) |
|  | Yes | 65 (6.9) | 20 (4.0) | 45 (10.0) |
| Years living in United States (N=958)<sup>1</sup> |  |  |  |  |
|  | <20 years | 451 (47.1) | 263 (52.3) | 188 (41.3) |
|  | ≥20 years | 507 (52.9) | 240 (47.7) | 267 (58.7) |
| Field worker occupation (N=1,089)<sup>1</sup> |  |  |  |  |
|  | No | 270 (24.8) | 134 (24.8) | 136 (24.8) |
|  | Yes | 819 (75.2) | 407 (75.2) | 412 (75.2) |
| Workplace setting (N=1,098)<sup>1</sup> |  |  |  |  |
|  | Indoor and outdoor | 260 (23.7) | 134 (24.4) | 126 (23) |
|  | Outdoor only | 838 (76.3) | 417 (75.6) | 422 (77) |
| Body mass index (N = 1,071)<sup>1</sup> |  |  |  |  |
|  | <18.5, underweight | 4 (0.4) | 2 (0.4) | 2 (0.4) |
|  | 18.5-24.9, normal | 186 (17.4) | 101 (19.0) | 85 (15.7) |
|  | 25-29.9, overweight | 409 (38.2) | 202 (38.0) | 207 (38.3) |
|  | 30-39.9, obese | 427 (39.9) | 205 (38.6) | 222 (41.1) |
|  | ≥ 40, severely obese | 45 (4.2) | 21 (4.0) | 24 (4.4) |
| Cigarette smoking (N=1,098)<sup>1</sup> |  |  |  |  |
|  | Never smoked | 891 (81.1) | 446 (81) | 445 (81) |
|  | Former smoker | 158 (14.4) | 86 (16) | 72 (13) |
|  | Current smoker | 49 (4.5) | 18 (3.3) | 31 (5.7) |
| Wearing a face covering when in proximity to others outside the home (N=1,099)<sup>1</sup> |  |  |  |  |
|  | Some of the time, rarely, or never | 86 (7.8) | 44 (8.0) | 42 (7.7) |
|  | All or most of the time | 1013 (92.2) | 507 (92.0) | 506 (92.3) |
| Regular handwashing after touching objects outside the home (N=1,099)<sup>1</sup> |  |  |  |  |
|  | Some of the time, rarely, or never | 33 (3.0) | 18 (3.3) | 15 (2.7) |
|  | All or most of the time | 1066 (97.0) | 533 (96.7) | 533 (97.3) |
| Housing type |  |  |  |  |
|  | House | 597 (46.5) | 330 (58.3) | 267 (37.2) |
|  | Apartment | 576 (44.9) | 193 (34.1) | 383 (53.4) |
|  | Trailer/mobile home | 45 (3.5) | 23 (4.1) | 22 (3.1) |
|  | Hotel/motel | 38 (3.0) | 8 (1.4) | 30 (4.2) |
|  | Other | 27 (2.1) | 12 (2.1) | 15 (2.1) |
| Household size (N=1,282) |  |  |  |  |
|  | 0-2 others | 222 (17.3) | 100 (17.7) | 122 (17.0) |
|  | 3-4 others | 557 (43.5) | 207 (36.6) | 350 (48.8) |
|  | ≥5 others | 503 (39.2) | 258 (45.7) | 245 (34.2) |
| Persons per bedroom (N=1,280) | 0-2 persons | 854 (66.7) | 337 (59.8) | 517 (72.2) |
|  | >2-4 persons | 387 (30.2) | 206 (36.7) | 181 (25.3) |
|  | >4 persons | 39 (3.0) | 21 (3.7) | 18 (2.5) |
| Child aged <5 years in household | No | 821 (64.0) | 345 (61.0) | 476 (66.4) |
|  | Yes | 462 (36.0) | 221 (39.0) | 241 (33.6) |
| SARS-CoV-2 infection (N=1,278) | Negative | 1,134 (88.7) | 469 (83.2) | 665 (93.1) |
|  | Positive | 144 (11.3) | 95 (16.8) | 49 (6.9) |

SARS-CoV-2: Severe acute respiratory syndrome coronavirus 2, as detected by clinical transcription-mediated amplification testing from oropharyngeal specimens. We indicate the total number of individuals providing answers to each question with missing data.
<sup>1</sup>Variable collected among primary study participants only and not available for participants in the expanded study. We present disaggregated data for the primary and supplemental study populations in **Table S1**.
<sup>2</sup>We detail characteristics of participants who reported speaking indigenous languages at home, versus other participants, in **Table S11**.

### Pneumococcal detection

We detected *lytA* in 342 samples (26.7% of 1,283; **Table 2**). Pneumococcal carriage (based on presence of *lytA* and *piaB*) was present in 117 samples (9.1%), and higher-density pneumococcal carriage (c*_T_<*35 for *lytA* and *piaB*) was present in 53 samples (4.1%). Prevalence of pneumococcal carriage was 11.1% (63/566) among participants recruited in clinical settings and 7.5% (54/717) among participants recruited through outreach testing. Prevalence of higher-density pneumococcal carriage was 5.5% (31/566) and 3.1% (22/717) in clinical and outreach venues, respectively, while prevalence of any *lytA*-positive oral Streptococcal carriage was 29.3% (166/566) and 24.5% (176/717) in clinical and outreach venues, respectively (**Table S2**). Restricting the sample to participants with negative SARS-CoV-2 testing results, prevalence of pneumococcal carriage, higher-density pneumococcal carriage, and *lytA*-positive oral Streptococcal carriage was 7.5% (85/1,134), 3.0% (34/1,134), and 24.6% (279/1,134), respectively, and did not differ appreciably according to recruitment setting (<1% prevalence differences among participants enrolled in clinical and community settings).

**Table 2:** Detection of pneumococcal carriage in the study population.

| Outcome | Participants, *n* (%) |  |  |
| --- | --- | --- | --- |
|  | All | Clinic | Outreach |
|  | *N*=1,283 | *N*=566 | *N*=717 |
| Any pneumococcal carriage | 117 (9.1) | 63 (11.1) | 54 (7.5) |
| Higher-density pneumococcal carriage | 53 (4.1) | 31 (5.5) | 22 (3.1) |
| Any oral streptococcal (*lytA*-positive) carriage | 342 (26.7) | 166 (26.3) | 176 (24.5) |
| *Sample restricted to individuals with negative SARS-CoV-2 test results* | *N*=1,134 | *N*=469 | *N*=665 |
| Any pneumococcal carriage | 85 (7.5) | 37 (7.9) | 48 (7.2) |
| Higher-density pneumococcal carriage | 34 (3.0) | 15 (3.2) | 19 (2.9) |
| Any oral streptococcal (*lytA*-positive) carriage | 279 (24.6) | 116 (24.7) | 163 (24.5) |

SARS-CoV-2: Severe acute respiratory syndrome coronavirus 2, as detected by clinical transcription-mediated amplification testing from oropharyngeal specimens. Pneumococcal detection was defined as $c_T$ values below 40 for both the *lytA* and *piaB* targets. Higher-density pneumococcal carriage was defined as $c_T$ values below 35 for both the *lytA* and *piaB* targets. Oral streptococcal carriage detection was defined as $c_T$ values below 40 for *lytA* target.

### Predictors of pneumococcal carriage

In comparison to non-carriers, pneumococcal carriers tended to be younger (OR=0.74 [0.62-0.89] for every increase by 10 years in age), more likely to speak indigenous languages in their households (OR=3.94 [2.36-6.60]), less likely to have completed high school (OR=0.48 [0.26-0.90]), and more likely to be engaged in field work tasks (OR=2.02 [1.12-3.63]); **Table 3**; **Table 4**). Pneumococcal carriers had also resided in the US for shorter periods of time than non-carriers (OR=0.75 [0.61-0.93] for every increase by 10 years), although this variable was inherently related to participant age.

**Table 3:**
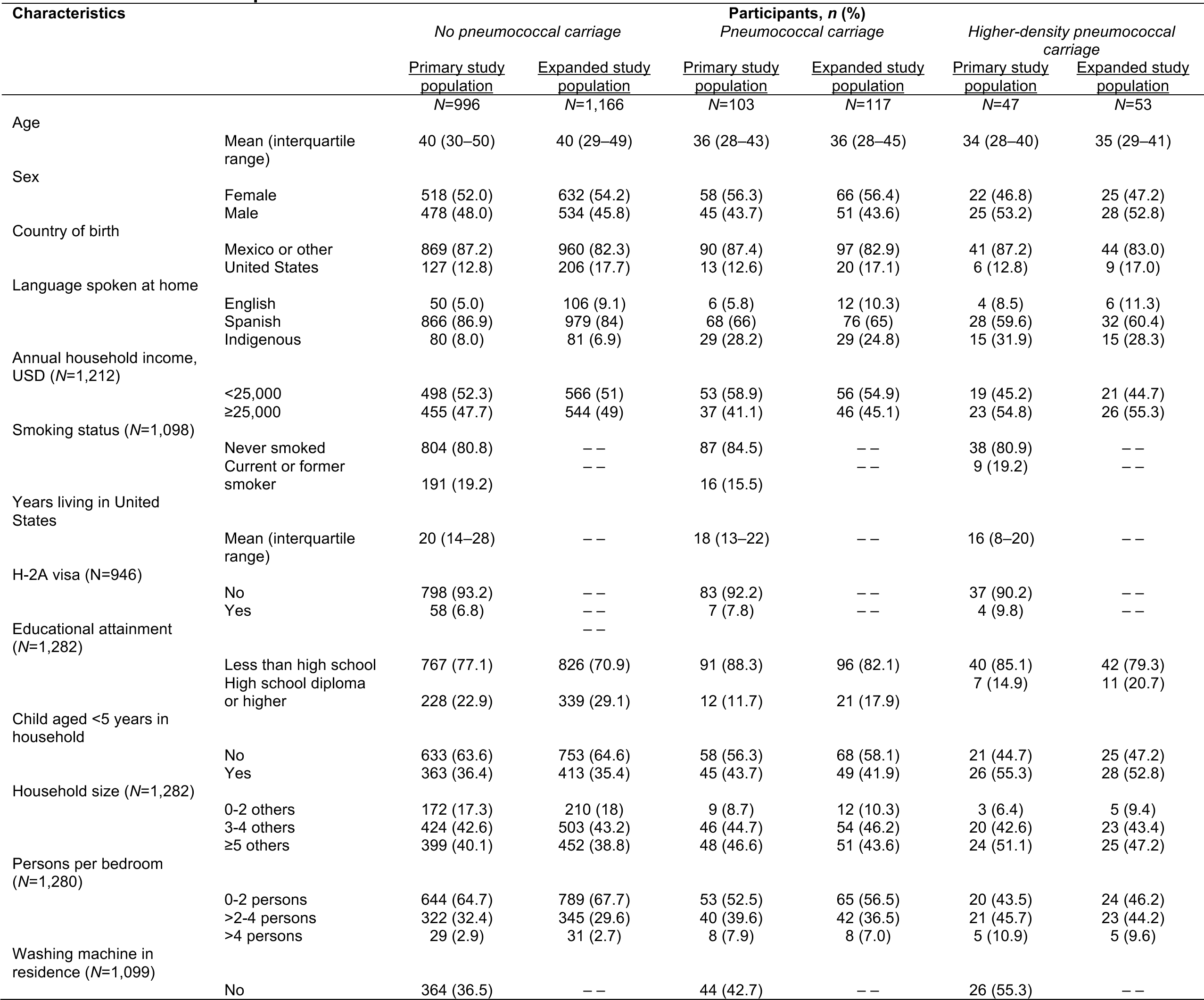
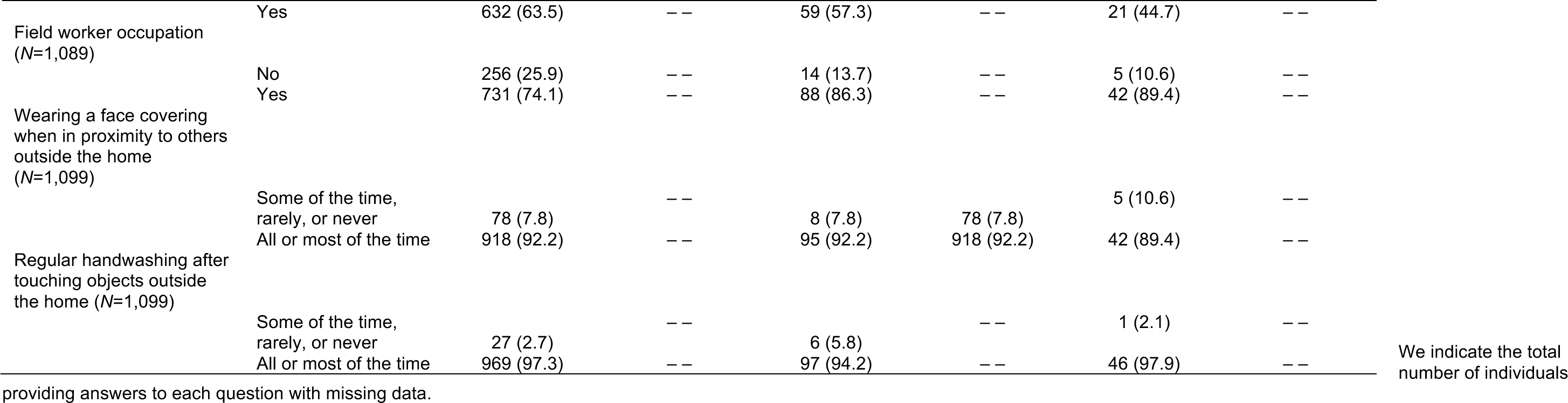
Characteristics of pneumococcal carriers and non-carriers.

**Table 4:**
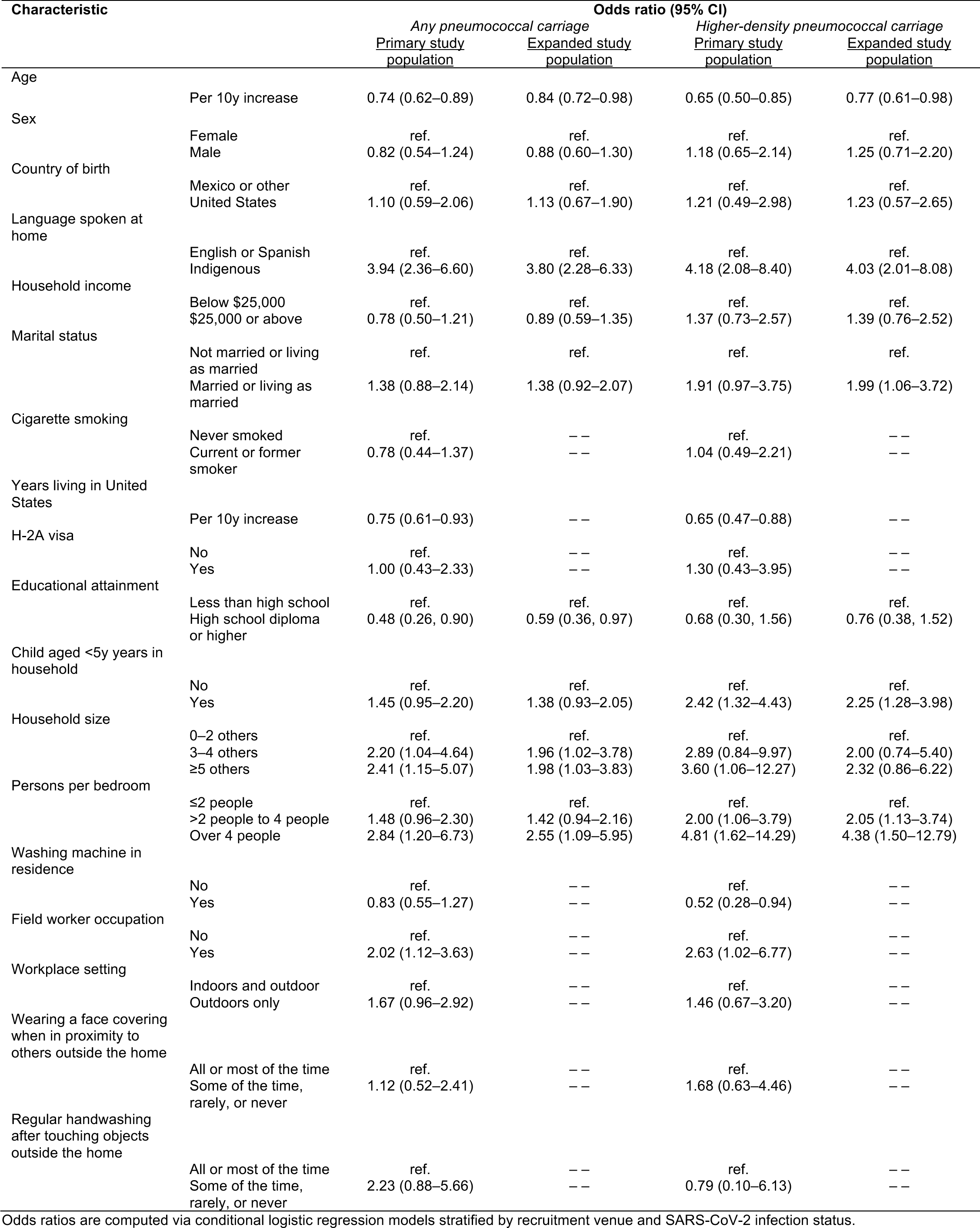
Associations of pneumococcal carriage and high-density carriage with participant characteristics.

Younger age and speaking an indigenous language at home also predicted higher-density pneumococcal carriage, along with a lack of access to in-home laundry facilities (OR=0.52 [0.28-0.94]).

With regard to household exposures, pneumococcal carriers were more likely than non-carriers to be exposed to a child aged <5 years at home (OR=1.45 [0.95-2.20]); to live in households with >2-4 persons per bedroom (OR=1.48 [0.96-2.30] vs. <2 persons per bedroom) or >4 persons per bedroom (OR=2.84 [1.20-6.73] vs. <2 persons per bedroom); and to live in households with 3-4 others (OR=2.20 [1.04-4.64] vs. <3 others) or ≥5 others (OR=2.41 [1.15-5.07] vs. <3 others; **Table 4**). For the same exposures, greater effect sizes were apparent for the outcome of higher-density pneumococcal carriage.

Associations between increased risk of pneumococcal carriage and exposure to crowding were also apparent in analyses restricted to participants who did not report exposure to a child aged <5 years at home (**Table 5**; **Table S3**). Among participants residing within households without a child aged <5 years, those living in households with >2 persons per bedroom had 2.05 (1.18-3.59) fold higher odds of pneumococcal carriage and 5.01 (1.96-12.83) fold greater odds of higher-density pneumococcal carriage in comparison to those living in households with ≤2 persons per bedroom.

**Table 5.** Associations of pneumococcal carriage and higher-density carriage with crowding among participants without residential exposure to children aged <5 years.

| Characteristic |  | Odds ratio (95% CI) |  |  |  |
| --- | --- | --- | --- | --- | --- |
|  |  | Any pneumococcal carriage |  | Higher-density pneumococcal carriage |  |
|  |  | Primary study population | Expanded study population | Primary study population | Expanded study population |
| Household size | 0–2 others | ref. | ref. | ref. | ref. |
|  | 3–4 others | 2.10 (0.93–4.72) | 1.95 (0.94–4.04) | 2.93 (0.63–13.66) | 2.22 (0.61–8.07) |
|  | ≥5 others | 2.18 (0.91–5.23) | 1.98 (0.89–4.41) | 3.81 (0.78–18.64) | 2.99 (0.78–11.51) |
|  | ≥3 others | 2.13 (0.98–4.63) | 1.96 (0.97–3.95) | 3.25 (0.73–14.37) | 2.48 (0.72–8.57) |
| Persons per bedroom | ≤2 persons | ref. | ref. | ref. | ref. |
|  | >2–4 persons | 1.78 (0.98–3.23) | 1.68 (0.95–2.97) | 4.41 (1.67–11.83) | 4.00 (1.64–9.74) |
|  | ≥4 persons | 2.93 (0.86–9.90) | 2.58 (0.77–8.59) | 6.49 (1.15–36.63) | 5.14 (0.94–28.00) |
|  | >2 persons | 2.05 (1.18–3.59) | 1.92 (1.13–3.25) | 5.01 (1.96–12.83) | 4.42 (1.90–10.28) |

Likewise, those living in households with ≥3 other inhabitants had 2.13 (0.98-4.63) fold higher odds of pneumococcal carriage and 3.25 (0.73-14.37) fold higher odds of higher-density pneumococcal carriage in comparison to those living in households with <3 others.

Factors independently associated with pneumococcal carriage in multivariate analyses included speaking an indigenous language (aOR=3.20 [1.81-5.62] for speakers of indigenous languages versus English or Spanish) and infrequent handwashing after contact with objects outside the household (OR=2.68 [1.04-6.91] for handwashing some of the time, rarely or never, versus all or most of the time; **Table 6**). The same risk factors were independently associated with any *lytA*-positive oral Streptococcal carriage (**Table S4**; **Table S5**). Additionally, higher-density pneumococcal carriage was independently associated with exposure to children aged <5 years at home (aOR=2.09 [1.12-3.92]) and with living in households with greater degrees of crowding (aOR=1.27 [0.99-1.61] per additional person per bedroom in the household). We obtained similar results in analyses restricted to participants testing negative for SARS-CoV-2 infection and in analyses excluding participants with any missing data (**Table S6, Table S7**).

**Table 6:**
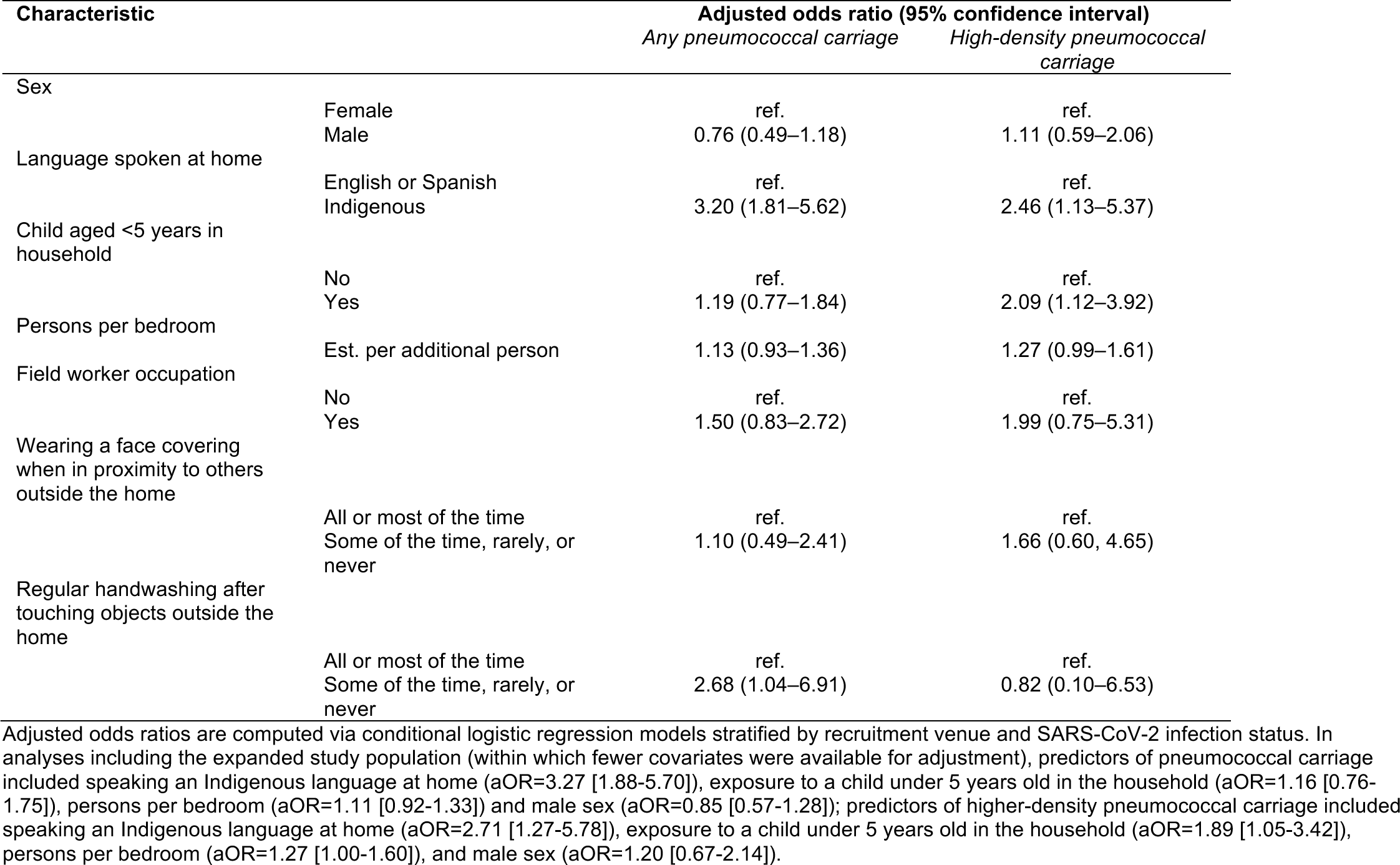
Adjusted associations of pneumococcal carriage and higher-density carriage with participant characteristics.

| Characteristic |  | Adjusted odds ratio (95% confidence interval) |  |
| --- | --- | --- | --- |
|  |  | *Any pneumococcal carriage* | *High-density pneumococcal carriage* |
| Sex |  |  |  |
|  | Female | ref. | ref. |
|  | Male | 0.76 (0.49–1.18) | 1.11 (0.59–2.06) |
| Language spoken at home |  |  |  |
|  | English or Spanish | ref. | ref. |
|  | Indigenous | 3.20 (1.81–5.62) | 2.46 (1.13–5.37) |
| Child aged <5 years in household |  |  |  |
|  | No | ref. | ref. |
|  | Yes | 1.19 (0.77–1.84) | 2.09 (1.12–3.92) |
| Persons per bedroom |  |  |  |
|  | Est. per additional person | 1.13 (0.93–1.36) | 1.27 (0.99–1.61) |
| Field worker occupation |  |  |  |
|  | No | ref. | ref. |
|  | Yes | 1.50 (0.83–2.72) | 1.99 (0.75–5.31) |
| Wearing a face covering when in proximity to others outside the home |  |  |  |
|  | All or most of the time | ref. | ref. |
|  | Some of the time, rarely, or never | 1.10 (0.49–2.41) | 1.66 (0.60, 4.65) |
| Regular handwashing after touching objects outside the home |  |  |  |
|  | All or most of the time | ref. | ref. |
|  | Some of the time, rarely, or never | 2.68 (1.04–6.91) | 0.82 (0.10–6.53) |

Adjusted odds ratios are computed via conditional logistic regression models stratified by recruitment venue and SARS-CoV-2 infection status. In analyses including the expanded study population (within which fewer covariates were available for adjustment), predictors of pneumococcal carriage included speaking an Indigenous language at home (aOR=3.27 [1.88-5.70]), exposure to a child under 5 years old in the household (aOR=1.16 [0.76-1.75]), persons per bedroom (aOR=1.11 [0.92-1.33]) and male sex (aOR=0.85 [0.57-1.28]); predictors of higher-density pneumococcal carriage included speaking an Indigenous language at home (aOR=2.71 [1.27-5.78]), exposure to a child under 5 years old in the household (aOR=1.89 [1.05-3.42]), persons per bedroom (aOR=1.27 [1.00-1.60]), and male sex (aOR=1.20 [0.67-2.14]).

### Pneumococcal carriage density

Among participants carrying pneumococci, those exposed to a child in the household aged <5 years, on average, had *piaB* c*_T_* values 2.04 (0.36-3.73) units lower than those not exposed to a child in the household aged <5 years (**Figure 1**, **Table S8**). Similarly, pneumococcal carriers exposed to household crowding (defined as >2 persons per bedroom in the household), on average, had *piaB* c*_T_* values 2.44 (0.80-4.11) units lower than those not exposed to household crowding. We did not identify any participant characteristics or exposures associated with differences in *lytA* c*_T_* values.

**Figure 1:**
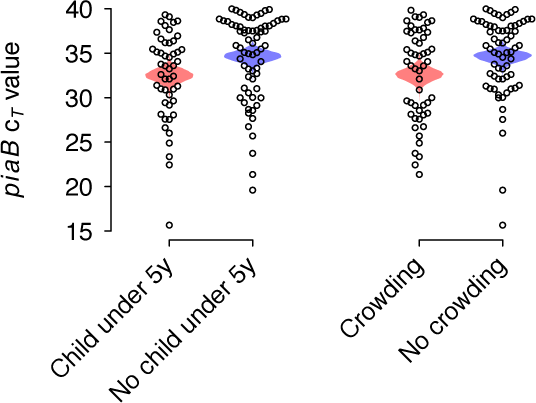
Factors predicting pneumococcal carriage density. We illustrate *piaB* c*_T_* value distributions among participants found to carry pneumococci based on their exposure to children aged <5 years in the household and to crowding in the household. Crowding is defined as living in a household with ≥2 persons per bedroom. Mean between-group differences in c*_T_* values were −2.04 (−33.73, −0.36) for households with or without children aged <5y and −2.44 (−34.11, −0.80) for crowded vs. uncrowded households (**Table S8**); including data from the expanded study population, mean between-group differences in c*_T_* values were −2.08 (−33.85, −0.33) for crowded vs. uncrowded households, and −2.17 (−33.95, −0.41) for households with or without children aged <5y. Shaded polygons illustrate the distributions of mean estimates within each stratum based on bootstrap resampling

### Serotype distribution

The most commonly detected serotypes among pneumococcal carriers were 15A (*n*=22, 18.8%), 24F/A/B (*n*=14, 12.0%), 10A (*n*=12, 10.3%), and 23B (*n*=10, 8.5%; **Table 7**). Among 78 participants with any serotype detected (66.7% of all 117 pneumococcal carriers), 13 (16.7%), 14 (17.9%), and 32 (41.0%) participants carried serotypes covered by PCV13, PCV15, and PCV20, respectively. Detection of a pneumococcal serotype was 33.2% (3.2-67.9%) more likely among pneumococcal carriers with concurrent SARS-CoV−2 infection than among those without concurrent SARS-CoV−2 infection (26/32 [81.3%] and 52/85 [61.2%], respectively), in line with our prior finding of higher pneumococcal carriage density among participants with concurrent SARS-CoV−2 infection [20]. Overall, serotype distributions did not differ appreciably among participants who were infected or not infected with SARS-CoV−2 according to the *χ*^2^ test, although point estimates suggested greater proportions of pneumococcal carriers with concurrent SARS-CoV−2 infection carried serotypes targeted by PCV13, PCV15, and PCV20 (overall, and among those with serotypes identified).

**Table 7.** Pneumococcal serotypes detected, stratified by SARS-CoV−2 infection status.

| Serotype |  | Participants, *n* (%) |  |  |
| --- | --- | --- | --- | --- |
|  |  | All participants | SARS-CoV-2 negative participants | SARS-CoV-2 positive participants |
|  |  | *N*=117 | *N*=85 | *N*=32 |
| 15A |  | 22 (18.8) | 15 (17.6) | 7 (21.9) |
| 24F/A/B |  | 14 (12) | 8 (9.4) | 6 (18.8) |
| 10A |  | 12 (10.3) | 10 (11.8) | 2 (6.3) |
| 23B |  | 10 (8.5) | 7 (8.2) | 3 (9.4) |
| 22A/F |  | 5 (4.3) | 3 (3.5) | 2 (6.3) |
|  | 22A | 3 (2.6) | 2 (2.4) | 1 (3.1) |
|  | 22F | 1 (0.9) | 0 (0) | 1 (3.1) |
|  | Indistinguishable 22A/F | 1 (0.9) | 1 (1.2) | 0 (0) |
| 9L/N |  | 5 (4.3) | 2 (2.4) | 3 (9.4) |
| 35F/47F |  | 5 (4.3) | 5 (5.9) | 0 (0) |
| 3 |  | 4 (3.4) | 1 (1.2) | 3 (9.4) |
| 34 |  | 4 (3.4) | 2 (2.4) | 2 (6.3) |
| 13 |  | 4 (3.4) | 4 (4.7) | 0 (0) |
| 28A/F |  | 4 (3.4) | 3 (3.5) | 1 (3.1) |
| 11A/D/E |  | 4 (3.4) | 2 (2.4) | 2 (6.3) |
| 14 |  | 3 (2.6) | 2 (2.4) | 1 (3.1) |
| 15B/C |  | 3 (2.6) | 2 (2.4) | 1 (3.1) |
| 23F |  | 2 (1.7) | 2 (2.4) | 0 (0) |
| 7F/A |  | 2 (1.7) | 1 (1.2) | 1 (3.1) |
| 16F |  | 1 (0.9) | 1 (1.2) | 0 (0) |
| 19A |  | 1 (0.9) | 0 (0) | 1 (3.1) |
| 6A/B |  | 1 (0.9) | 1 (1.2) | 0 (0) |
| 12F/44 |  | 0 (0) | 0 (0) | 0 (0) |
| 8 |  | 0 (0) | 0 (0) | 0 (0) |
| 38 |  | 0 (0) | 0 (0) | 0 (0) |
| 1 |  | 0 (0) | 0 (0) | 0 (0) |
| 20 |  | 0 (0) | 0 (0) | 0 (0) |
| Any serotype detected |  | 78 (66.7) | 52 (61.2) | 26 (81.3) |
| Serotypes grouped according to PCV inclusion |  |  |  |  |
|  | PCV13 serotypes | 13 (11.1) | 7 (8.2) | 6 (18.8) |
|  | PCV15 serotypes | 14 (12.0) | 7 (8.2) | 7 (21.9) |
|  | PCV20 (including both 15B/C) | 32 (27.4) | 21 (24.7) | 11 (34.4) |
| Number of serotypes detected per sample |  |  |  |  |
|  | 1 | 53 (45.3) | 34 (40.0) | 19 (59.4) |
|  | 2 | 20 (17.1) | 16 (18.8) | 4 (12.5) |
|  | 3 | 3 (2.6) | 1 (1.2) | 2 (6.3) |
|  | 4 | 2 (1.7) | 1 (1.2) | 1 (3.1) |

Serotype findings are presented only for specimens where serotype-specific $c_T$ values were within the range of $\pm 2$ from the *piaB* $c_T$ value. Serotypes 4, 9V/9A, 17F, 21, 23A, 33A/33F, and 35B are excluded due to low specificity of these probes in previous publications. Serogroup 6A/B was differentiated from 6A/B/C/D using conventional PCR methods.

### Symptoms

Among participants who were found not to be infected with SARS-CoV−2, odds of reporting any solicited symptoms or any respiratory symptoms did not differ in association with pneumococcal carriage detection or carriage density (**Table S9**; **Table S10**). However, higher-density pneumococcal carriers had greater adjusted odds of reporting fever (aOR=4.72 [1.27-17.45]), sweating (aOR=4.42 [1.21-16.14]), and difficulty breathing (aOR=9.16 [2.36-35.48]) in comparison to participants who did not carry pneumococci.

## DISCUSSION

Our findings provide insight into several facets of pneumococcal carriage among adults within a population experiencing low socioeconomic status and prevalent exposure to overcrowding. Within our study, 7.5% of participants without SARS-CoV−2 infection carried pneumococci, including 7.9% of those recruited in clinics and 7.2% of those recruited in community settings. Our findings implicate exposure to young children (aged <5 years) as a key risk factor for pneumococcal carriage, consistent with other studies enrolling either working-age or elderly adult samples [10,24,30]. However, we also found that exposure to household overcrowding (>2 persons per bedroom) was independently associated with increased risk of pneumococcal carriage, including among adults residing in households without young children. Each of these associations was strengthened when evaluating an outcome of higher-density pneumococcal carriage (*lytA* and *piaB* c*_T_*<35); moreover, residential exposure to children aged <5 years and living in overcrowded housing were each independently associated with increased carriage density among pneumococcal carriers. Because higher-density carriage can be a transient state resulting from recent pneumococcal acquisition [29], these findings may indicate that individuals with residential exposure to young children and overcrowding acquire pneumococci with greater frequency. Taken together, our findings suggest that in addition to transmission from young children to adults, transmission among adults may be an important feature of pneumococcal epidemiology in populations with prevalent exposure to residential overcrowding. This finding may help to explain observations such as the occurrence of outbreaks among otherwise healthy non-elderly adult populations living or working in close, congregate settings [11-17].

Our study also identified several risk factors for pneumococcal carriage related to lower socioeconomic status within this community, including indigenous ethnic or cultural background (indicated by speaking an indigenous language at home), lower educational attainment, and engagement in fieldwork tasks (which receive lower pay in comparison to other agricultural occupations such as machine operation or truck driving). Individuals without access to in-home laundry facilities were also at increased risk of higher-density carriage, which may either proxy similar socioeconomic status considerations or result from transmission within laundromats (*lavanderías*), which represent important community gathering places within the study setting and remained open during the pandemic. However, high degrees of collinearity among these variables, and their association with household crowding, constrain estimation of independent adjusted effects given the study’s limited sample size.

Because our study was undertaken during early phases of the COVID-19 pandemic, our questionnaires also addressed behaviors aiming to reduce pathogen transmission such as wearing face coverings and regular handwashing. Whereas use of face coverings was not associated with participants’ risk of pneumococcal carriage, we identified that participants who reported infrequent handwashing had higher adjusted odds of pneumococcal carriage and any *lytA*-positive oral Streptococcal carriage than those who reported handwashing with greater frequency. These findings may suggest differing transmission pathways for pneumococci—for instance, dependent upon direct interpersonal contact or contact with fomites—as compared to respiratory viruses, which are more prominently associated with airborne transmission [31]. Consistent with these findings, prior interventional studies have demonstrated that hygiene interventions reduced pneumococcal carriage prevalence in daycare centers [32] as well as the incidence of acute respiratory infections in such settings, despite a lack of similar effects against influenza transmission [33]. These distinct transmission dynamics may help to explain the persistence of pneumococcal carriage within communities amid implementation of nonpharmaceutical interventions that nearly eliminated transmission of many respiratory viruses during the COVID-19 pandemic [34]. Poor hand hygiene practices have also been implicated as a risk factor for upper respiratory carriage of bacteria such as *Haemophilus influenzae* and *Staphylococcus aureus* [35-37], which may share transmission pathways with pneumococcus.

Overall, 11.1%, 12.0%, and 27.4% of pneumococcal carriers in our study were found to carry serotypes targeted by PCV13, PCV15, and PCV20, respectively. These findings are broadly in agreement with prior evidence [38] that PCV20 (in comparison to PCV15) offers a greater improvement in coverage of serotypes prevalent in mucosal carriage or infections of the upper respiratory tract over PCV13. However, a majority of carried serotypes within our study were not covered by currently-available PCVs. While the relative pathogenicity of non-PCV20 serotypes remains to be established, this finding may underscore a persisting need among adults for next-generation PCV products offering broader coverage of serotypes relevant to adults.

Our study has several limitations. First, because exposures in our observational study could not be assigned through randomization, we cannot rule out the possibility that unobserved confounding variables may impact measured associations. Socioeconomic status is likely a key confounder due to its potential association with exposure to residential overcrowding and may be imperfectly measured by variables such as indigenous ethnicity, education, income, and fieldwork tasks available in our study. Second, our design is cross-sectional in nature, and thus does not offer an opportunity to establish temporality between exposures and pneumococcal carriage acquisition. Longitudinal studies sampling household members and other close contacts may provide further insight into the impact of identified risk factors on transmission dynamics. Third, exposures of interest such as overcrowding were prevalent within our study population; studies enrolling more diverse participants (including individuals with higher socioeconomic status) may be positioned to identify greater effect sizes due to greater variation in such exposure. Fourth, we did not collect information about recent antibiotic receipt, which may have relevance for pneumococcal carriage detection and density outcomes, especially among individuals experiencing symptomatic illness. Fifth, stronger associations of measured risk factors with higher-density pneumococcal carriage may relate to the greater specificity of this outcome in comparison to pneumococcal carriage detection at the *lytA* and *piaB* c*_T_*<40 threshold. While it is also possible that viral infections besides SARS-CoV−2 contribute to higher-density pneumococcal carriage, the low degrees of circulation of other respiratory viruses during the summer and fall of 2020 [39] make it unlikely that such infections would influence findings in our specific study context. Last, the infeasibility of colony-based pneumococcal methods for assessment of adult carriage limited our ability to characterize serotype distributions in this study population. While restricting serotyping results to samples with c*_T_* values within ±2 of values observed for *piaB* helped to maximize specificity, we could not study all serotypes due to the nonspecific association of some *cps* genes with other colonizing bacteria. Regardless, the proportion of samples containing pneumococcus for which we were able to identify ≥1 serotype was consistent with other studies using similar molecular methods [24,40].

In conclusion, our study identifies exposure to young children and residential overcrowding—including in households without young children—as important risk factors for pneumococcal carriage among adults in low-income agricultural communities within California. The association of these factors with higher carriage density further suggests individuals encountering young children or crowded conditions within the residential environment acquire pneumococcal carriage with greater frequency. While confirming prior understanding that young children serve as a reservoir for transmission of pneumococci, these findings also suggest that transmission among adults may play an important feature of pneumococcal epidemiology among populations exposed to overcrowding. Strategies to prevent transmission under such conditions, or to reduce individuals’ risk of severe pneumococcal disease through vaccination, may be of value for certain populations.<colcnt=3>

## Data Availability

Data produced in the present study can be made available upon reasonable request to the authors with signed data use agreements in place.

## ACKNOWLEDGMENTS

This work was supported by the Innovative Genomics Institute at the University of California, Berkeley and by Pfizer, Inc. (grant 61775823 to JAL).

## CONFLICTS OF INTEREST

JAL discloses receipt of grant funding from Pfizer, Inc. and Merck, Sharp & Dohme, and consulting honoraria from Pfizer, Inc., Merck, Sharp & Dohme, and VaxCyte, Inc. ALW discloses receipt of grant funding from Pfizer, Inc., Merck, Sharp & Dohme, SalivaDirect, Inc., Valvi.io, and Shield T3, and consulting honoraria from Pfizer, Inc., Merck, Sharp & Dohme, Diasorin, PPS Health, Co-Diagnostics, and Global Diagnostic Systems. All other authors declare no competing interests.

## Supplemental laboratory methods

### Sample processing and culture enrichment

Participants were instructed to provide the saliva that naturally pools in the mouth. These saliva specimens were frozen at –20°C and then stored at –80°C until transport to laboratory. Samples were thawed on ice after arrival at the laboratory; after thawing, 100µL of unprocessed saliva was spread onto trypticase soy agar supplemented with 7% sheep’s blood and 5mg/L gentamicin. Plates were then incubated at 37°C with 5% CO2 for 14-19 hours. Bacterial growth was harvested into 2.1mL of 10% glycerol brain-heart infusion broth (BHI), and stored at –20°C. The remaining raw saliva not used for culture enrichment was stored at –80°C.

### Isolation of bacterial DNA

Culture-enriched samples were thawed and vortexed then transferred into sterile 1.5mL Eppendorf tubes prefilled with lysis buffer containing 40mg/mL lysozyme and 75U/mol mutanolysin. Next, the mixture was incubated for 90 minutes at 37°C, followed by the addition of 20µL of Proteinase K and another incubation of 60 minutes at 56 °C. Subsequent extraction steps used the QIAGEN DNeasy Blood and Tissue Kit (QIAGEN; Hilden, Germany) following manufacturer’s instructions. DNA was eluted into 50µL elution buffer and nucleic acid concentration was measured using a NanoDrop spectrophotometer. Samples with a DNA concentration <20ng/µL underwent a second extraction. Experimental control DNA was extracted with the biolate method from pure culture. Briefly, colonies grown overnight on a TSA-GENT plate at 37°C with 5% CO2 were extracted and suspended in 60µL nuclease-free water. The sample was then placed into a water bath heated to 100°C and boiled for 10 minutes before being centrifuged for 10 minutes at 14,000 rpm. The supernatant was removed, and nucleic acid concentration was measured. Positive and negative control DNA was diluted with nuclease-free water to a concentration of 50ng/µL. All extracted DNA samples were stored at –20°C.

### Molecular detection of *S. pneumoniae*

Culture-enriched DNA was tested via quantitative PCR (qPCR) for presence of the *lytA* and *piaB* genes to determine pneumococcal presence (defined as identification with 40 or fewer cycle threshold values [c*_T_*] for both genes). Gene detection via qPCR was conducted with primer concentrations at 400nM and 300nM, and probe concentrations at 75nM and 200nM, for lytA and piaB, respectively. All qPCR assays tested 2.5µL of DNA template in 25µL reactions. Thermal cycling conditions were as follows: 3 minutes at 95°C for initial denaturation; 15 seconds at 98°C for denaturation; and 30 seconds at 60°C for annealing. Positive controls consisted of a diluted series of reference strain ATCC 49619. We used DNA from human saliva previously characterized as negative for *S. pneumoniae* and DNA from *E. coli* reference strain ATCC 25922 as our negative controls.

---

**Table S1.** Characteristics of the primary and expanded study populations.

| Characteristics |  | Participants, n (%) |  |  |
| --- | --- | --- | --- | --- |
|  |  | All<br>N=1,283 | Primary population<br>N=1,099 | Supplemental population<br>N=184 |
| Recruitment venue | Clinical setting | 566 (44) | 551 (50) | 15 (8) |
|  | Outreach setting | 717 (56) | 548 (50) | 169 (92) |
| Age range, years | 18-29 | 344 (27) | 274 (25) | 70 (38) |
|  | 30-39 | 306 (24) | 272 (25) | 34 (18) |
|  | 40-49 | 327 (25) | 290 (26) | 37 (20) |
|  | ≥50 | 306 (24) | 263 (24) | 43 (23) |
| Sex | Female | 698 (54) | 576 (52) | 122 (66) |
|  | Male | 585 (46) | 523 (48) | 62 (34) |
| Country of birth | Mexico | 1,012 (79) | 917 (83) | 86 (47) |
|  | United States | 226 (18) | 140 (13) | 95 (52) |
|  | Other | 45 (3.5) | 42 (3.8) | 3 (2) |
| Language spoken at home | Spanish | 1,055 (82) | 934 (85) | 62 (34) |
|  | English | 118 (9) | 56 (5) | 121 (66) |
|  | Indigenous language | 110 (89) | 109 (10) | 1 (<1) |
| Education | Never attended school | 63 (5) | 62 (6) | 1 (<1) |
|  | Some primary school | 460 (36) | 422 (38) | 38 (21) |
|  | Primary school completed | 251 (20) | 235 (21) | 16 (9) |
|  | Some high school | 148 (12) | 139 (13) | 9 (5) |
|  | High school completed | 360 (28) | 240 (22) | 120 (65) |
| Annual household income, USD (N=1,212) | <25,000 | 622 (51) | 551 (53) | 71 (42) |
|  | 25,000-34,999 | 284 (23) | 257 (25) | 27 (16) |
|  | 35,000-49,000 | 195 (16) | 159 (15) | 36 (21) |
|  | ≥ 50,000 | 111 (9) | 76 (7) | 35 (21) |
| Housing type | House | 597 (47) | 520 (47) | 77 (42) |
|  | Apartment | 576 (45) | 474 (43) | 102 (55) |
|  | Trailer/mobile home | 45 (4) | 43 (4) | 2 (1) |
|  | Hotel/motel | 38 (3) | 37 (3) | 1 (<1) |
|  | Other | 27 (2) | 25 (2) | 2 (1) |
| Community of residence | Greenfield | 309 (24) | 309 (28) | 0 (0) |
|  | Salinas | 485 (38) | 485 (44) | 0 (0) |
|  | Other | 489 (38) | 305 (28) | 184 (100) |
| Pneumococcal detection | *lytA* c<sub>T</sub> <40 | 342 (27) | 302 (27) | 40 (22) |
|  | *lytA* c<sub>T</sub> <40 and *piaB* c<sub>T</sub> <40 | 118 (9) | 104 (10) | 14 (18) |
|  | *lytA* c<sub>T</sub> <35 and *piaB* c<sub>T</sub> <35 | 59 (5) | 51 (5) | 8 (4) |

**Table S2:**
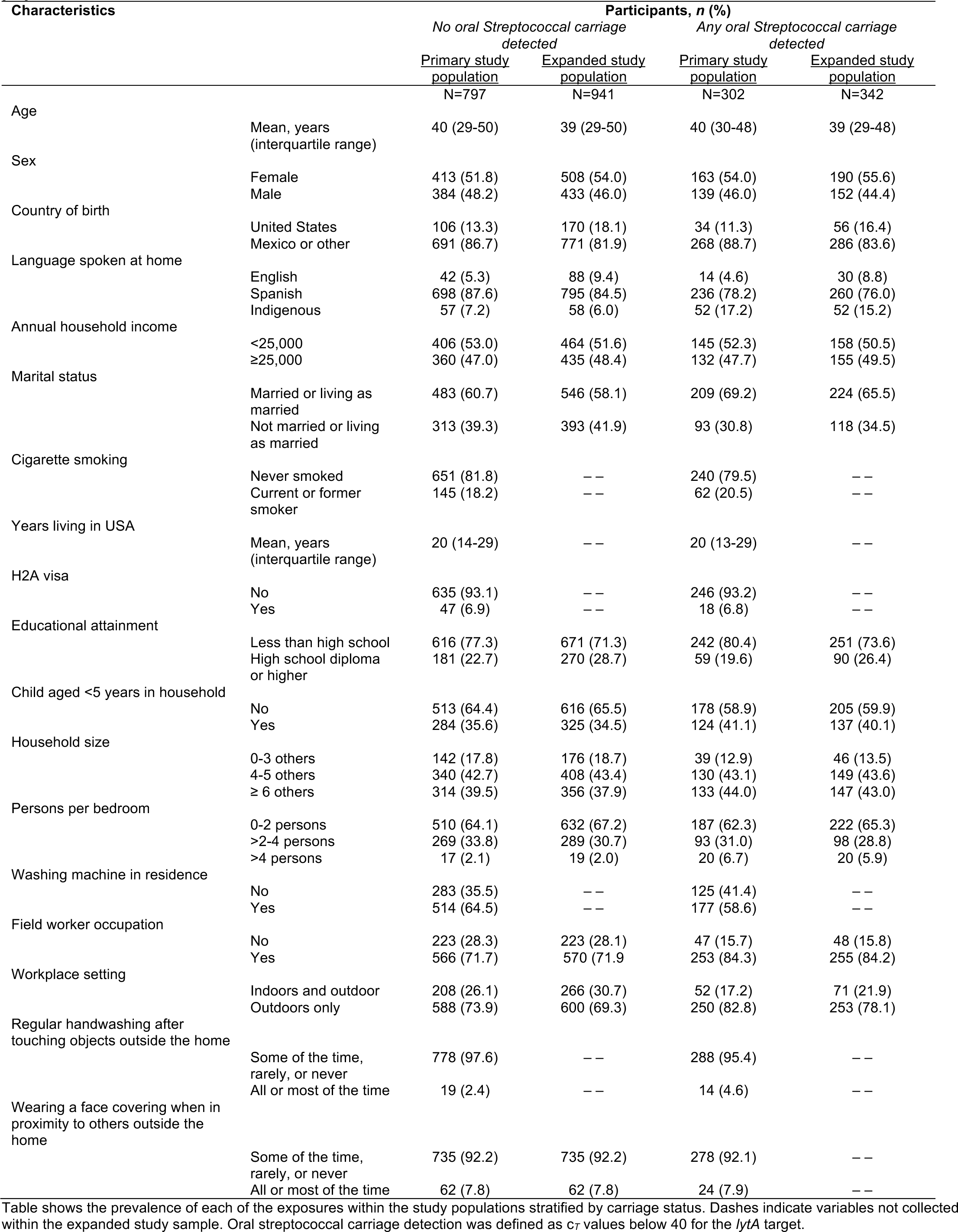
**Detection of other *lytA*-positive oral Streptococcal carriage within the primary and expanded study populations.**

**Table S3:**
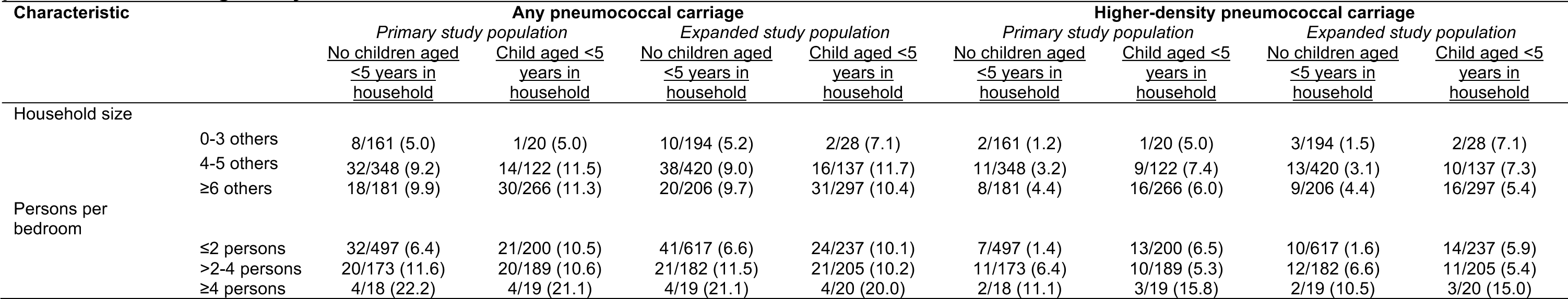
Detection of pneumococcal carriage among participants exposed to varying levels of household crowding, stratified according to the presence of a child aged <5 years in the household.

**Table S4:**
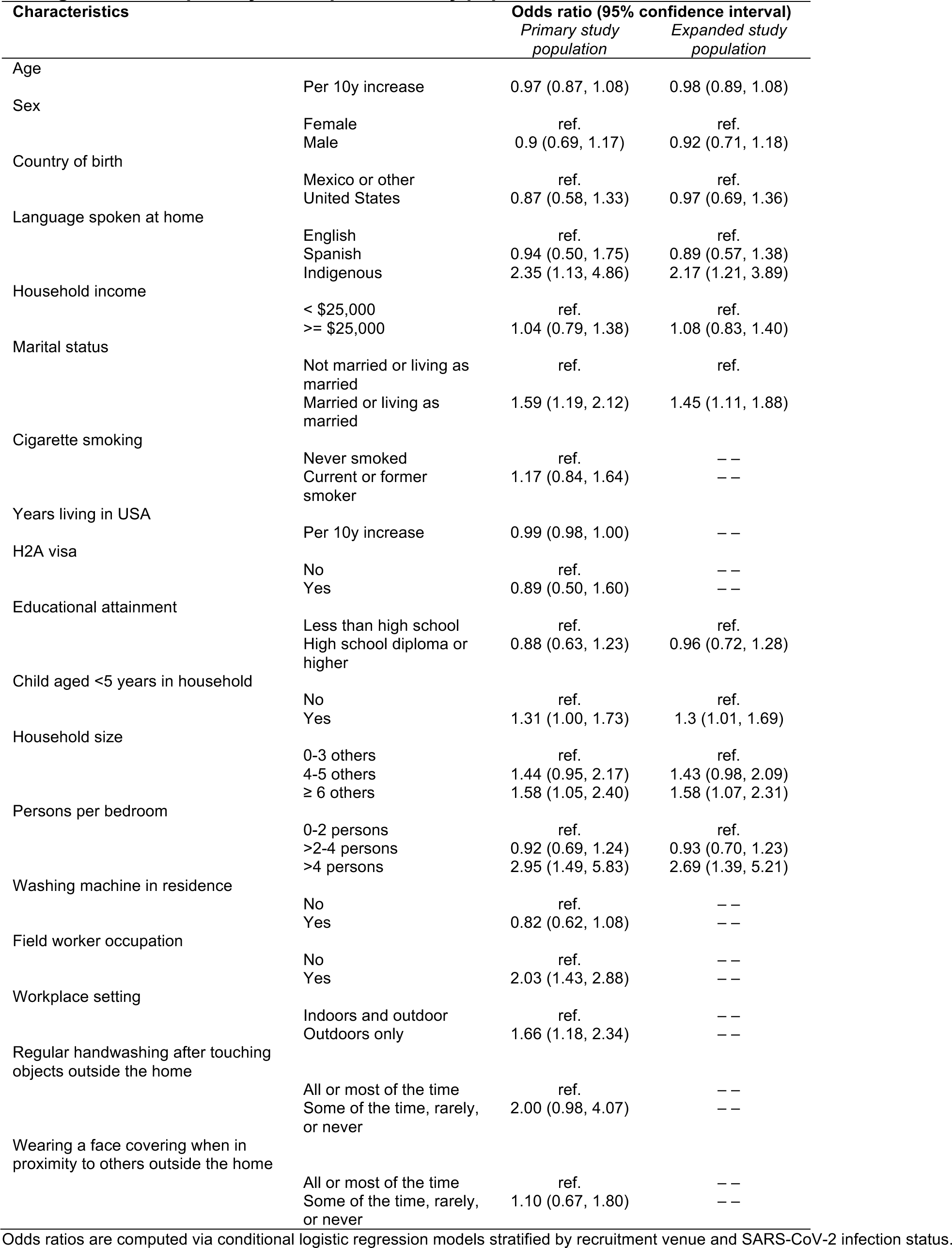
Associations of participant characteristics with detection of other *lytA*-positive oral Streptococcal carriage within the primary and expanded study populations.

**Table S5:** Adjusted association of exposures with other *lytA*-positive oral Streptococcal carriage within the primary study population.

| Characteristic |  | Adjusted odds ratio (95% confidence interval) |
| --- | --- | --- |
| Language spoken at home | English or Spanish | ref. |
|  | Indigenous | 1.97 (1.26, 3.09) |
| Child aged <5 years in household | No | ref. |
|  | Yes | 1.18 (0.89, 1.57) |
| Persons per bedroom | Fold change per additional person | 1.08 (0.94, 1.23) |
| Field worker occupation | No | ref. |
|  | Yes | 1.78 (1.25, 2.53) |
| Sex | Female | ref. |
|  | Male | 0.88 (0.67, 1.16) |
| Wearing a face covering when in proximity to others outside the home | All or most of the time | ref. |
|  | Some of the time, rarely, or never | 1.04 (0.63, 1.73) |
| Regular handwashing after touching objects outside the home | All or most of the time | ref. |
|  | Some of the time, rarely, or never | 2.10 (1.01, 4.34) |

Adjusted odds ratios are computed via conditional logistic regression models stratified by recruitment venue and SARS-CoV-2 infection status. In analyses including the expanded study population, speaking an Indigenous language at home (aOR=2.16 [1.38-3.36]) was a significant predictor of carriage.

**Table S6:** Adjusted association of exposures with pneumococcal carriage within the primary study population, excluding participants with any missing data.

| Characteristic |  | Adjusted odds ratio (95% confidence interval) |  |
| --- | --- | --- | --- |
|  |  | *Pneumococcal carriage* | *Higher-density pneumococcal carriage* |
| Language | English or Spanish | ref. | ref. |
|  | Indigenous | 2.92 (1.61, 5.29) | 2.30 (1.03, 5.12) |
| Child aged <5 years in household | No | ref. | ref. |
|  | Yes | 1.24 (0.80, 1.95) | 2.21 (1.17, 4.19) |
| Persons per bedroom | Est. per additional person | 1.14 (0.94, 1.38) | 1.28 (1.00, 1.63) |
| Field worker occupation | No | ref. | ref. |
|  | Yes | 1.54 (0.84, 2.82) | 1.95 (0.73, 5.18) |
| Sex | Female | ref. | ref. |
|  | Male | 0.74 (0.48, 1.15) | 1.08 (0.57, 2.03) |
| Regular handwashing after touching objects outside the home | All or most of the time | ref. | ref. |
|  | Some of the time, rarely, or never | 2.66 (1.03, 6.88) | 0.80 (0.10, 6.43) |
| Wearing a face covering when in proximity to others outside the home | All or most of the time | ref. | ref. |
|  | Some of the time, rarely, or never | 1.12 (0.51, 2.49) | 1.72 (0.62, 4.79) |

Adjusted odds ratios are computed via conditional logistic regression models stratified by recruitment venue and SARS-CoV-2 infection status. In analyses including the expanded study population, the significant predictor of pneumococcal carriage was speaking an Indigenous language at home (aOR=3.03 [1.72-5.36]); predictors of higher-density pneumococcal carriage included speaking an Indigenous language at home (aOR=2.50 [1.15-5.43]), household exposure to a child aged <5 years (aOR=2.0 [1.11-3.63]), and persons per bedroom within the household (aOR=1.28 [1.01-1.62]).

**Table S7:**
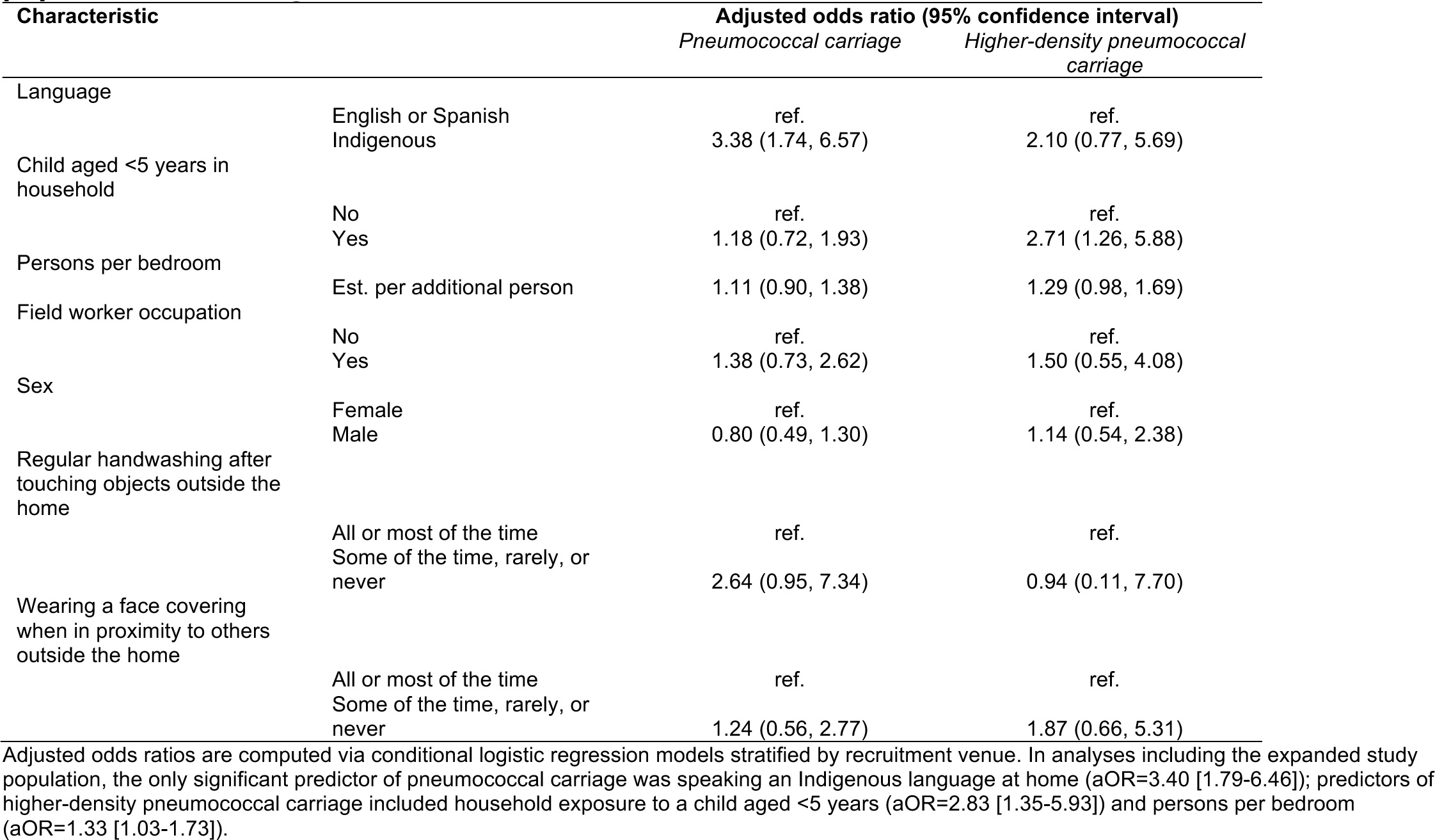
Adjusted association of exposures with pneumococcal carriage within the primary study population, excluding participants with SARS-CoV−2 infection.

| Characteristic |  | Adjusted odds ratio (95% confidence interval) |  |
| --- | --- | --- | --- |
|  |  | *Pneumococcal carriage* | *Higher-density pneumococcal carriage* |
| Language | English or Spanish | ref. | ref. |
|  | Indigenous | 3.38 (1.74, 6.57) | 2.10 (0.77, 5.69) |
| Child aged <5 years in household | No | ref. | ref. |
|  | Yes | 1.18 (0.72, 1.93) | 2.71 (1.26, 5.88) |
| Persons per bedroom | Est. per additional person | 1.11 (0.90, 1.38) | 1.29 (0.98, 1.69) |
| Field worker occupation | No | ref. | ref. |
|  | Yes | 1.38 (0.73, 2.62) | 1.50 (0.55, 4.08) |
| Sex | Female | ref. | ref. |
|  | Male | 0.80 (0.49, 1.30) | 1.14 (0.54, 2.38) |
| Regular handwashing after touching objects outside the home | All or most of the time | ref. | ref. |
|  | Some of the time, rarely, or never | 2.64 (0.95, 7.34) | 0.94 (0.11, 7.70) |
| Wearing a face covering when in proximity to others outside the home | All or most of the time | ref. | ref. |
|  | Some of the time, rarely, or never | 1.24 (0.56, 2.77) | 1.87 (0.66, 5.31) |

Adjusted odds ratios are computed via conditional logistic regression models stratified by recruitment venue. In analyses including the expanded study population, the only significant predictor of pneumococcal carriage was speaking an Indigenous language at home (aOR=3.40 [1.79-6.46]); predictors of higher-density pneumococcal carriage included household exposure to a child aged <5 years (aOR=2.83 [1.35-5.93]) and persons per bedroom (aOR=1.33 [1.03-1.73]).

**Table S8:** Association of participant characteristics with *lytA* and *piaB* c*_T_* values among pneumococcal carriers.

| Characteristic |  | Target: *lytA* |  | Target: *piaB* |  |
| --- | --- | --- | --- | --- | --- |
|  |  | Mean difference<br>(95% CI) | Adjusted mean<br>Difference (95% CI) | Mean difference<br>(95% CI) | Adjusted mean<br>difference (95% CI) |
| Sex |  |  |  |  |  |
|  | Female | ref. | ref. | ref. | ref. |
|  | Male | -0.50 (-2.44, 1.42) | -0.10 (-2.08, 1.86) | -1.51 (-3.22, 0.19) | -1.46 (-3.12, 0.22) |
| Language spoken at home |  |  |  |  |  |
|  | English or Spanish | ref. | ref. | ref. | ref. |
|  | Indigenous | 0.06 (-2.08, 2.18) | 0.00 (-2.31, 2.25) | -0.64 (-2.56, 1.25) | 0.94 (-1.01, 2.85) |
| Child aged <5 years in household |  |  |  |  |  |
|  | No | ref. | ref. | ref. | ref. |
|  | Yes | 1.22 (-0.71, 3.13) | 1.53 (-0.51, 3.56) | -2.04 (-3.73, -0.36) | -2.23 (-3.95, -0.49) |
| Persons per bedroom |  |  |  |  |  |
|  | 0-2 people | ref. | ref. | ref. | ref. |
|  | >2 people | -0.18 (-2.10, 1.73) | -0.57 (-2.64, 1.50) | -2.44 (-4.11, -0.80) | -2.19 (-3.94, -0.43) |
| Field worker occupation |  |  |  |  |  |
|  | No | ref. | ref. | ref. | ref. |
|  | Yes | 1.48 (0.96–2.30) | 0.54 (-2.41, 3.49) | 2.00 (1.06–3.79) | -0.62 (-3.09, 1.88) |
| SARS-CoV-2 infection |  |  |  |  |  |
|  | Negative | ref. | ref. | ref. | ref. |
|  | Positive | 0.52 (-1.67, 2.68) | 0.35 (-1.91, 2.59) | -1.24 (-3.18, 0.69) | -1.45 (-3.35, 0.45) |
| Regular handwashing after touching objects outside the home |  |  |  |  |  |
|  | All or most of the time | ref. | ref. | ref. | ref. |
|  | Some of the time, rarely, or never | 3.25 (-0.81, 7.28) | 4.41 (0.17, 8.62) | 1.30 (-2.38, 4.95) | 1.91 (-1.68, 5.49) |
| Wearing a face covering when in proximity to others outside the home |  |  |  |  |  |
|  | All or most of the time | ref. | ref. | ref. | ref. |
|  | Some of the time, rarely, or never | -2.91 (-6.46, 0.62) | -4.10 (-7.87, -0.33) | -1.53 (-4.75, 1.65) | -1.80 (-5.00, 1.37) |

SARS-CoV-2: Severe acute respiratory syndrome coronavirus 2, as detected by clinical transcription-mediated amplification testing from oropharyngeal specimens. In analyses including data from the expanded study population, there were no significant mean between-group differences in *lytA* c<sub>T</sub> values. Significant mean between-group differences (MD) in *piaB* c<sub>T</sub> values were apparent for crowded vs. uncrowded households (MD= -2.08 [-3.85, -0.33]; aMD -2.01 [-3.86, -0.19]) and households with or without children aged <5 years (MD = -2.17 [-3.95, -0.41]; aMD= -2.32 [-4.10, -0.53]).

**Table S9.**
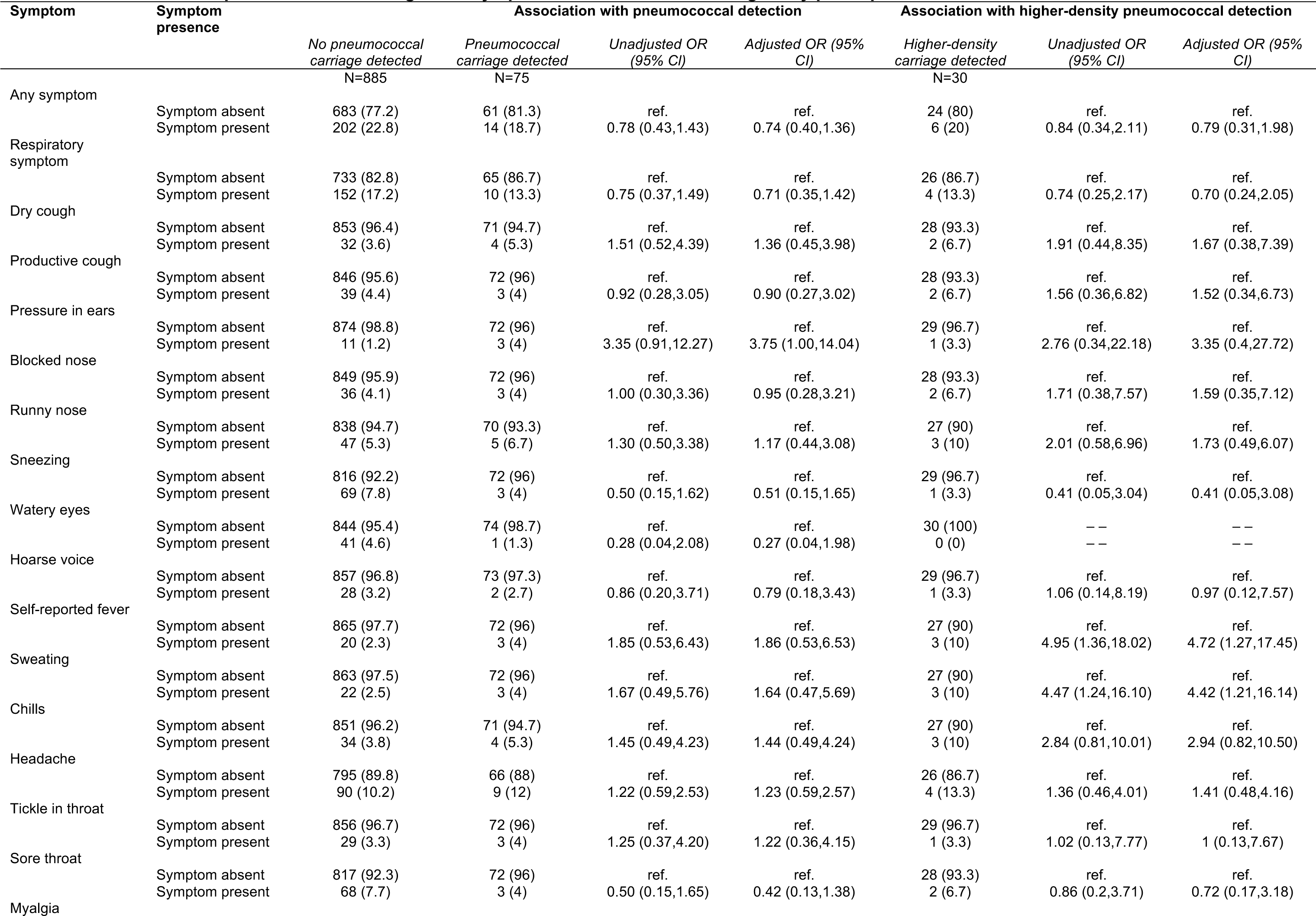
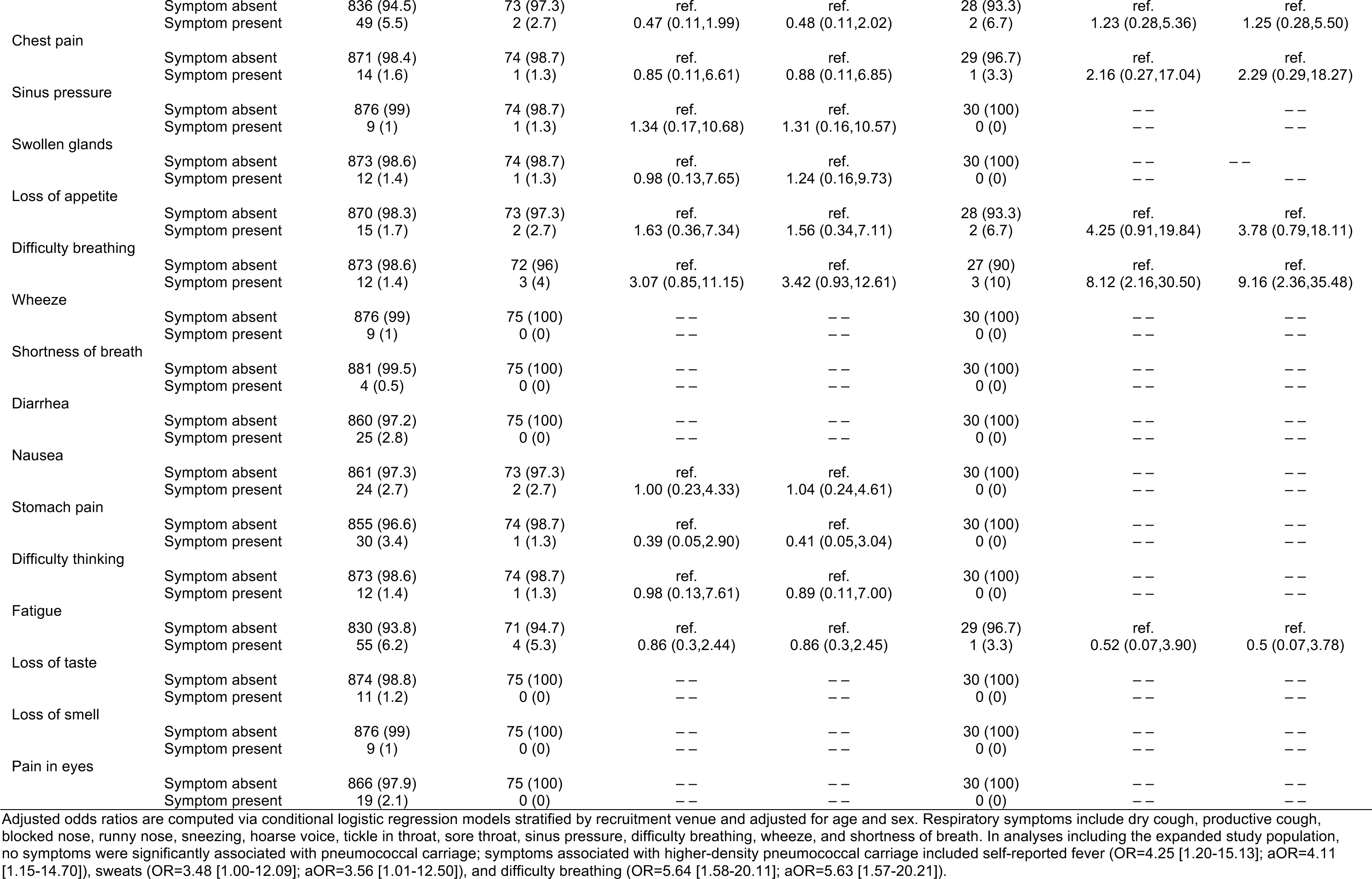
Associations of pneumococcal carriage with symptoms in last 2 weeks among study participants without SARS-CoV−2 infection.

**Table S10:**
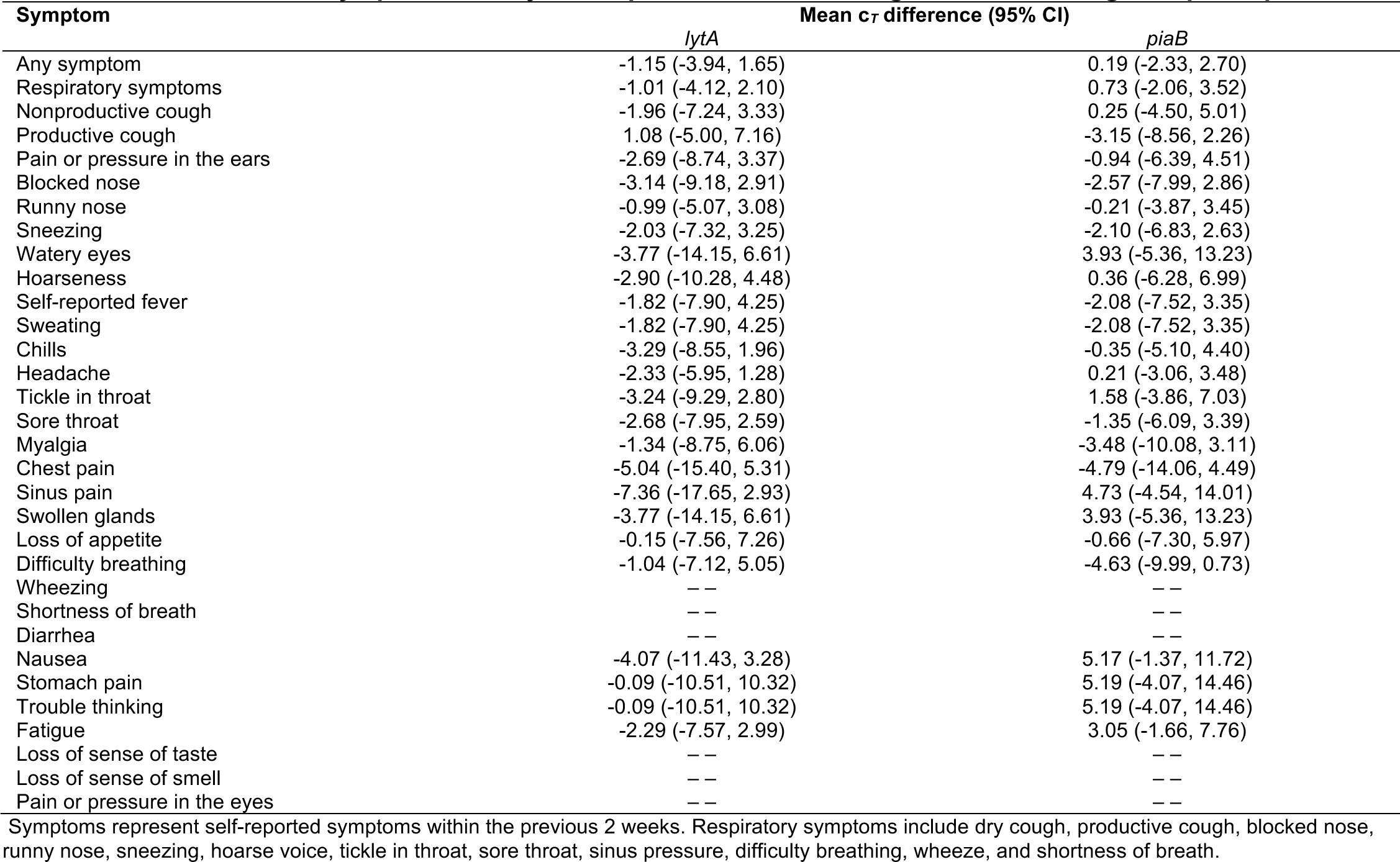
Association of symptoms with *lytA* and *piaB* c*_T_* values, among SARS-CoV−2 negative participants.

| Symptom | Mean $c_T$ difference (95% CI) | |
| --- | --- | --- |
|  | *lytA* | *piaB* |
| Any symptom | -1.15 (-3.94, 1.65) | 0.19 (-2.33, 2.70) |
| Respiratory symptoms | -1.01 (-4.12, 2.10) | 0.73 (-2.06, 3.52) |
| Nonproductive cough | -1.96 (-7.24, 3.33) | 0.25 (-4.50, 5.01) |
| Productive cough | 1.08 (-5.00, 7.16) | -3.15 (-8.56, 2.26) |
| Pain or pressure in the ears | -2.69 (-8.74, 3.37) | -0.94 (-6.39, 4.51) |
| Blocked nose | -3.14 (-9.18, 2.91) | -2.57 (-7.99, 2.86) |
| Runny nose | -0.99 (-5.07, 3.08) | -0.21 (-3.87, 3.45) |
| Sneezing | -2.03 (-7.32, 3.25) | -2.10 (-6.83, 2.63) |
| Watery eyes | -3.77 (-14.15, 6.61) | 3.93 (-5.36, 13.23) |
| Hoarseness | -2.90 (-10.28, 4.48) | 0.36 (-6.28, 6.99) |
| Self-reported fever | -1.82 (-7.90, 4.25) | -2.08 (-7.52, 3.35) |
| Sweating | -1.82 (-7.90, 4.25) | -2.08 (-7.52, 3.35) |
| Chills | -3.29 (-8.55, 1.96) | -0.35 (-5.10, 4.40) |
| Headache | -2.33 (-5.95, 1.28) | 0.21 (-3.06, 3.48) |
| Tickle in throat | -3.24 (-9.29, 2.80) | 1.58 (-3.86, 7.03) |
| Sore throat | -2.68 (-7.95, 2.59) | -1.35 (-6.09, 3.39) |
| Myalgia | -1.34 (-8.75, 6.06) | -3.48 (-10.08, 3.11) |
| Chest pain | -5.04 (-15.40, 5.31) | -4.79 (-14.06, 4.49) |
| Sinus pain | -7.36 (-17.65, 2.93) | 4.73 (-4.54, 14.01) |
| Swollen glands | -3.77 (-14.15, 6.61) | 3.93 (-5.36, 13.23) |
| Loss of appetite | -0.15 (-7.56, 7.26) | -0.66 (-7.30, 5.97) |
| Difficulty breathing | -1.04 (-7.12, 5.05) | -4.63 (-9.99, 0.73) |
| Wheezing | -- | -- |
| Shortness of breath | -- | -- |
| Diarrhea | -- | -- |
| Nausea | -4.07 (-11.43, 3.28) | 5.17 (-1.37, 11.72) |
| Stomach pain | -0.09 (-10.51, 10.32) | 5.19 (-4.07, 14.46) |
| Trouble thinking | -0.09 (-10.51, 10.32) | 5.19 (-4.07, 14.46) |
| Fatigue | -2.29 (-7.57, 2.99) | 3.05 (-1.66, 7.76) |
| Loss of sense of taste | -- | -- |
| Loss of sense of smell | -- | -- |
| Pain or pressure in the eyes | -- | -- |

Symptoms represent self-reported symptoms within the previous 2 weeks. Respiratory symptoms include dry cough, productive cough, blocked nose, runny nose, sneezing, hoarse voice, tickle in throat, sore throat, sinus pressure, difficulty breathing, wheeze, and shortness of breath.

**Table S11:**
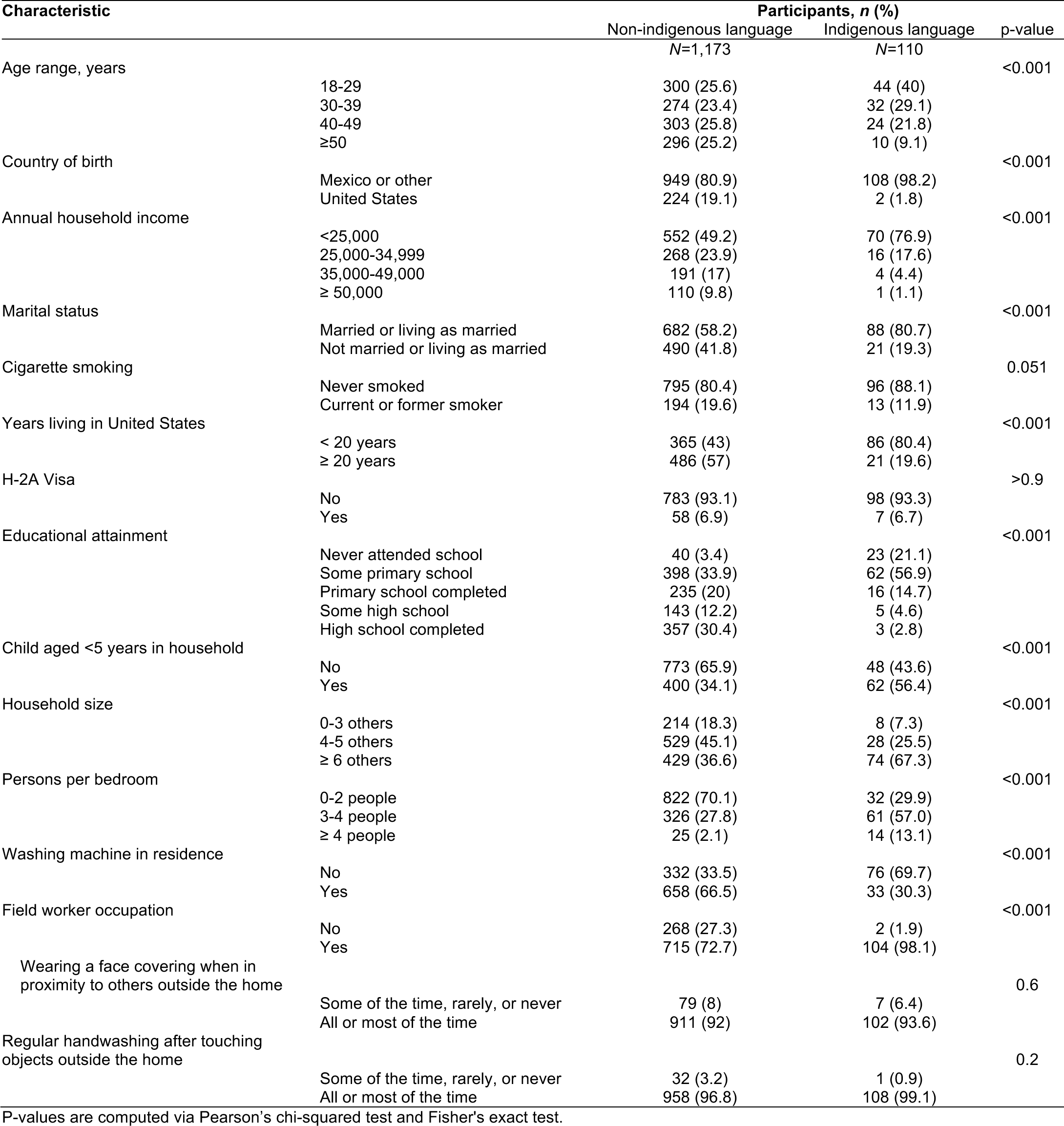
Distribution of risk factors among participants who reported speaking Indigenous languages at home and participants who did not report speaking indigenous languages.

## Notes

### Author Declarations

The Biomedical Human Subjects Committee of the Office for Protection of Human Subjects of the University of California, Berkeley gave ethical approval for this work.

